# An investigation of the effect of sex on resolution of the inflammatory response in healthy volunteers: protocol of the Resolve-Sex study

**DOI:** 10.1101/2025.09.12.25335580

**Authors:** Andrew Sullivan, Rayomand S. Khambata, Manikandan Subramanian, Asad Shabbir, Gianmichele Massimo, Nicki Dyson, Vikas Kapil, Thomas Godec, Annastazia Learoyd, Krishnaraj S. Rathod, Amrita Ahluwalia

## Abstract

**Background:** Inflammation is key in the initiation and progression of coronary artery disease (CAD). To date clinical trials testing strategies to limit inflammation have been limited by unwanted effects. Inflammatory resolution represents the active process of restoring tissue homeostasis following a pro-inflammatory response and offers a potential novel approach to limiting the inflammatory response by accelerating resolution, rather than dampening the host-defence response. Women with CAD have higher rates of morbidity and mortality compared to their male counterparts and sex differences exist in presentations, infarct patterns and plaque characteristics. The underlying reasons for these differences are not well understood. Differences in biology, in particular inflammation and the resolution of inflammation, are likely to play an important role. Our previous work has demonstrated that healthy females are more adept at resolving inflammation compared to males, although the exact mechanisms for this were unclear. In this study, we will explore the molecular mechanisms underlying resolution and particularly why this process is accelerated in females.

**Methods:** In this comparative, single centre study we will recruit 34 healthy volunteers (17 females and 17 males). A blister model of acute inflammation will be used with cantharidin applied over 3 consecutive days and blisters harvested on the fourth day. Blister fluid from 24h, 48h and 72h timepoints will be collected and analysed using several laboratory techniques including fluorophore labelled flow cytometry and cytokine analysis. The primary endpoint for the study is the comparison of the numbers of volunteers with blisters present at each time point between the sexes. In addition, blister volume, cell count and cells per ml of blister fluid at each time point will be compared between the sexes. Secondary endpoints include comparison of blister leukocyte subsets, cell death, apoptotic cells numbers, markers of blister efferocytosis, blister lactate and LDH levels. The study has a power of 80% to assess the primary endpoint.

**Discussion:** In this study we aim to identify the mechanisms involved in sex differences in the resolution of the acute inflammatory response in healthy volunteers.

**Trial registration:** ClinicalTrials.gov NCT05597098. The study was approved by the Yorkshire & The Humber – Bradford Leeds Research Ethics Committee, reference 22/YH/0244.

## Background

Coronary artery disease (CAD) remains the number one cause of death worldwide despite advances in treatment (1). Pre-menopausal women are relatively protected against CAD, however, after the menopause, this protection is lost and women experience increasing CAD with age, whereas in men the incidence peaks at approximately 70 years (2). Concerning data has emerged demonstrating that females experience worse morbidity and mortality following acute coronary syndromes (ACS) and revascularization compared to their male counterparts (3,4). Whilst this can be, in part, accounted for by disparities in medical care, even after accounting for this, differences remain (5–8). Furthermore, sex differences also exist in clinical presentations, infarct patterns and coronary plaque characteristics in CAD (5,9,10). It is likely that differences in biology are important in accounting for these differences, however, our understanding of the sex-based differences in the physiology of CAD are limited, with pre-clinical and clinical studies over the years primarily focused on male disease (11).

Inflammation is a key process in the initiation and progression of CAD. The success of therapeutic strategies aimed at attenuating inflammation have been limited by on-target but unwanted effects, specifically immunosuppression (12,13). Inflammatory resolution is an active process that restores tissue homeostasis following an inflammatory trigger and the consequent coordinated response to clear the trigger. This may offer the potential to target inflammation without the unwanted effects (14). Efferocytosis (the engulfment of apoptotic cells by phagocytic leucocytes) is considered a key requirement for resolution to occur (15). In vitro data and pathological studies have previously shown that inflammatory resolution and efferocytosis may be impaired in CAD and therefore this offers a potential novel therapeutic target (16). However, it is important first to understand the molecular mechanisms involved in health. Our previous data has shown that healthy females appear to be more adept at resolving inflammation compared to males using a cantharidin-induced blister model of acute inflammation. In that study, healthy females exhibited statistically significant enhanced blister resolution, reductions in blister volume and inflammatory monocyte numbers. This was associated with reduced activation state in all major leucocytes in female blisters and a trend towards higher levels of the pro-resolving mediators CXCL8 and IL-10 (17). The cantharidin blister model is a validated in vivo model of inflammation which is safe for study participants. In this study we will explore the molecular mechanisms involved in this phenomenon using the same model of acute inflammation, with a particular focus on efferocytosis. This will form the basis for further work to understand whether the process of inflammation resolution is impaired in CAD and whether deficiencies in this might underlie CAD pathogenesis in women who suffer from this and also women’s increased morbidity and mortality compared to men following an ACS.

## Methods and design

### Trial objectives

The principal research objective is to determine which step in the resolution of inflammation pathway, underlies differences between females and males. We also wish to determine whether neutrophil and monocyte subsets, efferocytosis and cell death pathways differ between females and males.

### Participant Selection

In this single-centre study based at Barts and the London Faculty of Medicine and Dentistry, Queen Mary University of London, UK, healthy volunteers will be recruited from Queen Mary University of London and from the Barts Health NHS Trust catchment area.

### Original Hypothesis

We hypothesise that women will exhibit enhanced inflammatory resolution compared to males and that this phenomenon will be exhibited by enhancement in the molecular pathways involved in inflammatory resolution, in particular efferocytosis.

### Primary Endpoints

1. Comparison of the numbers of volunteers with a blister present at each timepoint over 24-72h between the sexes.
2. Comparison of blister volume at each timepoint over 24-72h between the sexes.
3. Comparison of blister cell number at each timepoint over 24-72h between the sexes.

### Secondary endpoints

1. Comparison of blister leucocyte subsets (neutrophil and monocyte) between the sexes at each timepoint.
2. Comparison of blister lactate levels and lactate dehydrogenase (LDH) between the sexes at each timepoint.
3. Comparison of cell death, necrotic and apoptotic cell numbers between the sexes at each timepoint.
4. Comparison of markers of blister efferocytosis between the sexes at each timepoint.

### Inclusion Criteria

1. Healthy male and female volunteers.
2. Aged 18-45.
3. Volunteers who are willing to sign the consent form.

### Exclusion Criteria

1. Healthy subjects unwilling to consent.
2. Pregnant, or any possibility that a subject may be pregnant unless in the latter case a pregnancy test is performed with a negative result.
3. Current breast feeding.
4. History of any serious illnesses, including recent infections or trauma.
5. Subjects taking systemic medication (other than contraceptives).
6. Subjects with recent (2 weeks) or current antibiotic use.
7. Current smoking.
8. Subjects with any history of a blood-borne infectious disease such as Hepatitis B or C virus, or HIV.

### Study Design

This is a single centre, randomised healthy volunteer, comparative study taking place at Barts and the London Faculty of Medicine and Dentistry, Queen Mary University of London, UK. 34 healthy volunteers will be recruited (17 females and 17 males, aged 18-45 years). At the screening visit eligibility will be confirmed as per the inclusion and exclusion criteria and if eligible informed consent obtained by a clinician (see model consent form in supplementary documents). Figure 1 shows the study scheme and Figure 2 shows the SPIRIT figure summary. Screening will take place virtually or in person. Participants will then be assigned a randomisation number. Following randomisation, the participant is still eligible to undergo usual care should they require it during the study including concomitant planned and unplanned medical care (unrelated to the study itself) and medication for the time period of physical visits and biological sampling; such an event would exclude them from the study.

**Figure 1.**
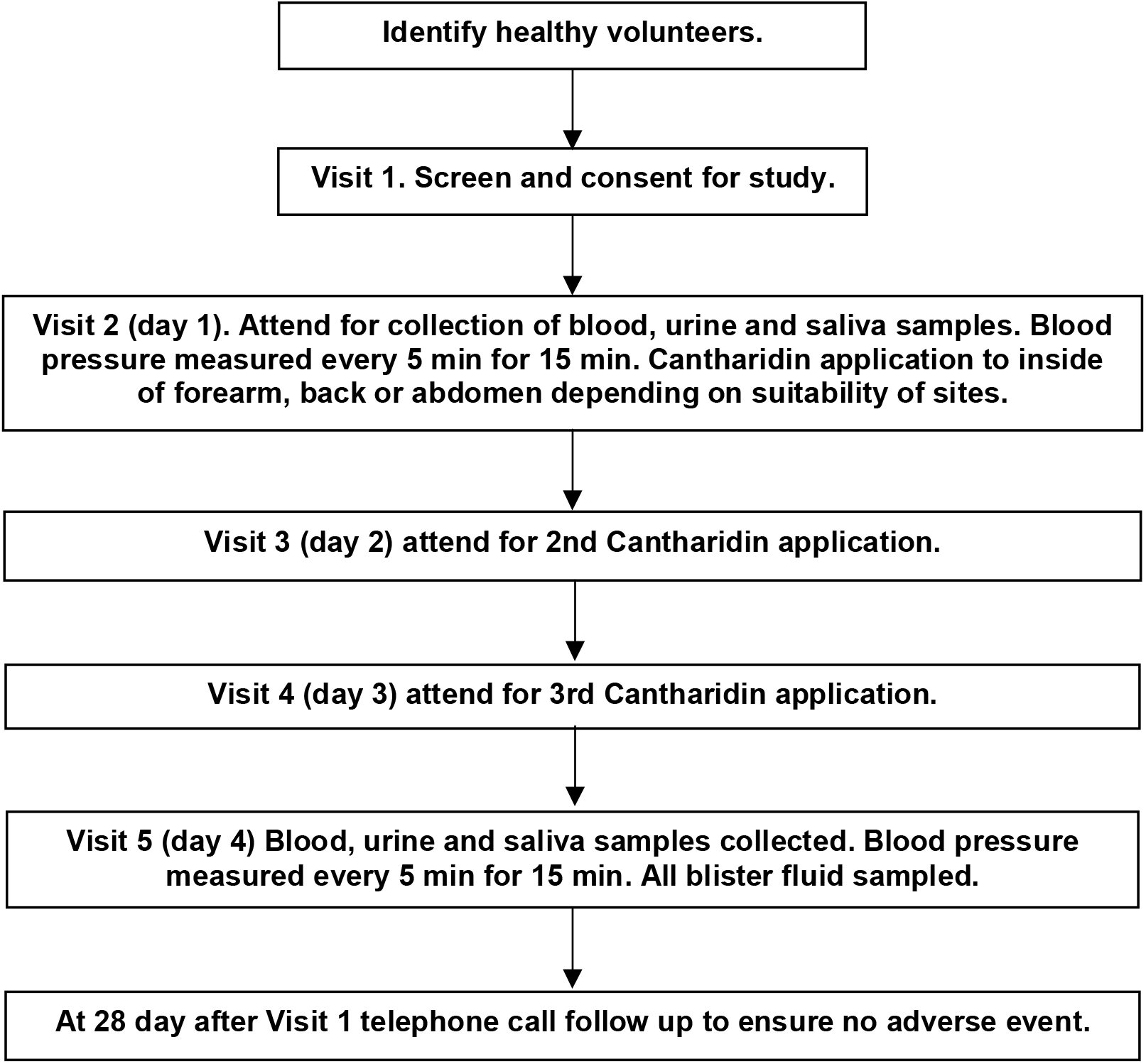
Resolve-Sex study scheme diagram.

**Figure 2.**
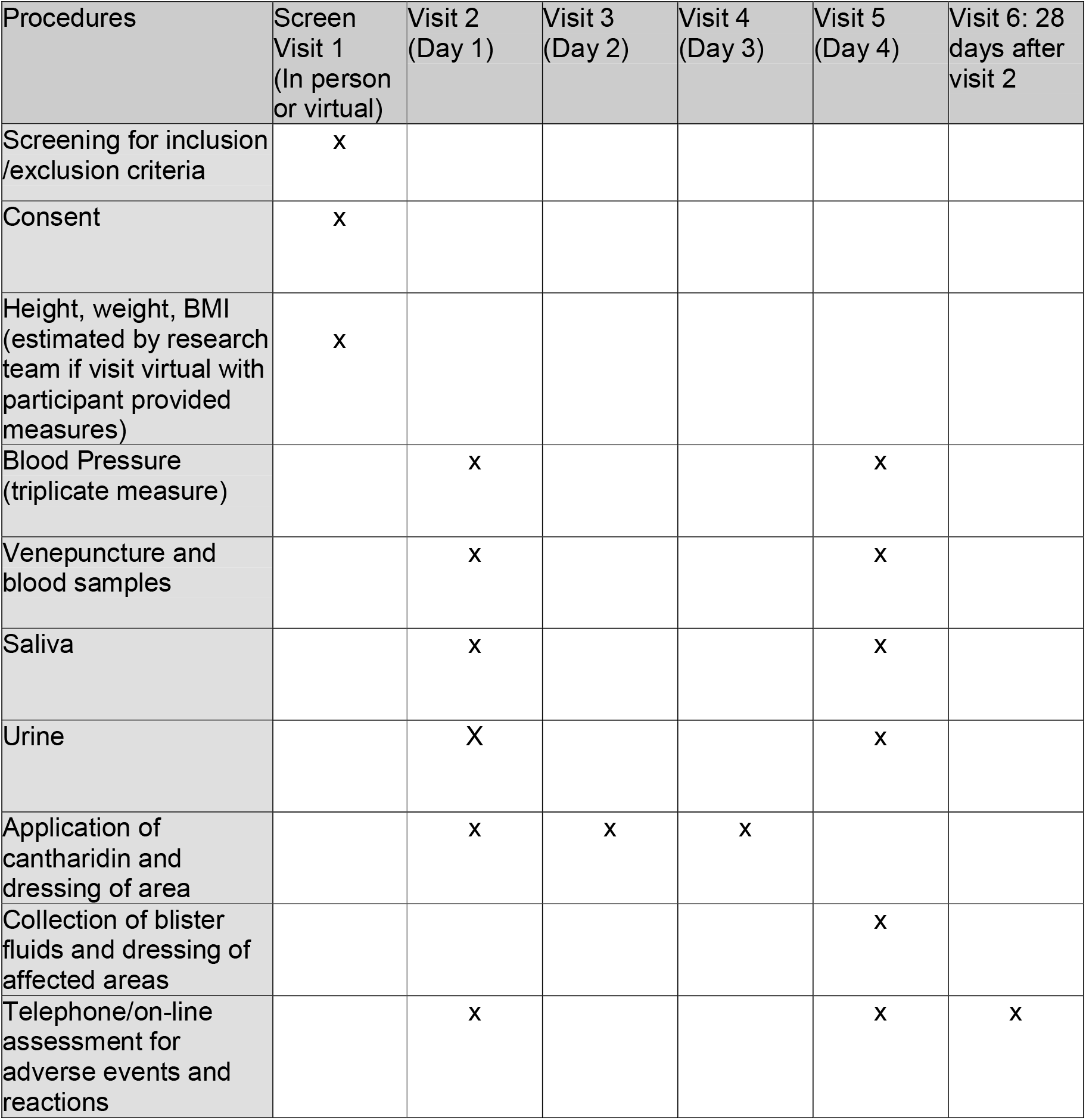
Summary spirit figure showing visit structure and events during the Resolve-Sex study.

At **visit 2**, At 9am ±2 h blood pressure will be measured every 5min for 15 min, a blood, urine and saliva sample will be collected, following which, a single application of cantharidin will be applied to the ventral aspect of the forearm, the back or the abdomen (by trained physician) as per the volunteer’s wishes.

At 24h later (**visit 3**) the subject will return and have cantharidin applied to a separate area, no less than 5cm away from the initial application on visit 1.

At **visit 4** 48h after Visit 2 the subject will return and have cantharidin applied to a separate area, no less than 5cm away from the first two applications.

At **visit 5** (72h after initial cantharidin application), blood pressure will be measured every 5min for 15 min.

A photograph of the participants’ blister application and appearance will be taken on a dedicated laboratory camera. The area included in the photograph will be minimised to include only the area limited to where the 3 blisters are e.g. the forearms. Therefore, it will not be possible to identify the participant from the photographs. Blister fluid will be sampled from blisters and a blood, urine and saliva sample will be collected. The blister area will be dressed with simple gauze and the participant advised to keep the blister roof intact for as long as possible to aid healing.

**Visit 6**: We will conduct a telephone interview 28 days following visit 1 to ascertain any adverse reactions or events.

### Randomisation and blinding process

This study cannot be blind since group allocation is pre-determined and based upon participant stated sex. However, objective analyses of blister volume, flow cytometry cell subtype, biochemical analysis of mediators of resolution can be assessed blind. Analysis of all data will be performed blind to sex as volunteers will be assigned a recruitment number and all samples/relevant data will be labelled with this unique number. The position of the blisters (24h, 48h, 72h) will be randomised to left upper, left lower and right upper sections of the forearms (or abdomen or back). A random number sequence of 1-3 will be used to determine the position for each participant with the number 1-3 relating to a pre-determined position on the forearm (or abdomen or back). The random number sequence was generated for the pre-specified sample size for each potential participant number and participants were allocated to a participant number in time order of enrolment. This was performed by a member of the research team.

### Study start and end dates

Recruitment commenced on 2 December 2022. The provisional end date of the study is December 2027.

## Methods to be used

### Blood samples

Blood samples (~5ml each) will be taken during the study as indicated above for:

- Haematological indices – leucocyte differential, platelet count, erythrocyte sedimentation rate.
- Biochemical indices – C-reactive protein, urate, U&E, oestrogen, FSH, testosterone.

These samples will be sent to the pathology laboratories at Barts Health NHS Trust and will be analysed as per routine laboratory methods.

Other plasma parameters to be measured included (chilled EDTA sample) – NO_3_^-^/NO_2_^-^ via chemiluminescence, cGMP levels, inflammatory mediators using commercially available cytokine/chemokine array or commercially available ELISAs. This sample is immediately centrifuged at 1300*g* at 4°C for 10 minutes. Plasma is separated, aliquoted with a cohort of samples incubated with isobutylmethylxanthine (IBMX, 100 μM) for cGMP measurements and stored at −80°C until measurement of biochemical parameters undertaken.

Blood samples will also be subject to immune profile analysis as described below.

### Urine samples

Clean-catch mid-stream urine samples will be collected into sterile gallipots and aliquoted into eppendorfs and stored at −80°C until measurement of NO_3_ ^-^ and NO_2_ ^-^ are undertaken by chemiluminescence.

### Saliva samples

Subjects will be instructed to allow saliva to drip from their tongue/lower lip into a sterile gallipot. Saliva will be aliquoted into 1.5mL microcentrifuge tubes and some spun at 14000rpm at 4°C for 10 minutes and the supernatant separated and stored at −80°C until measurement of NO_3_ ^-^ and NO_2_ ^-^ are undertaken by chemiluminescence. One sample will be collected directly into an Oragene DNA isolation vial for later DNA extraction. This analysis will be performed on anonymised samples therefore it will not be possible to link the results back to the subject. It therefore will not reveal any hidden medical diagnoses of subjects.

### Cantharidin-induced blister formation and fluid collection

10μL cantharidin (cantharone 0.1%, Dormer Laboratories, Toronto, Canada) will be applied to filter-paper discs on the ventral aspects of the forearm, the back or the leg as previously described and be dressed appropriately (17–21). Blister fluid will be harvested at 24h (acute phase) 48h (start of resolution) and 72h (resolution phase) after the acute inflammatory stimulus is applied, by carefully piercing the side of the blister with a 25G needle and rolling a pipette tip over the surface of the blister to express blister fluid, which is collected in a siliconised pipette and the fluid is stored on ice for further analysis.

### Inflammatory cell and cytokine analysis

Blood and fluid collected from the blister will be analysed by standard laboratory techniques including fluorophore labelled flow cytometry (CD45, CD66b, CD14, CD16, CD62L, CXCR4, CD206, CCR7, MERTK, Annexin V, Zombie for viability) and ELISA to measure inflammatory cytokines/chemokines including IL-10. Remaining inflammatory cells from blister fluid as well as peripheral blood mononuclear cells and polymorphonuclear neutrophils isolated from blood will subsequently be subject to bulk RNA extraction using the Qiagen RNeasy Mini Kit according to the manufacturer’s instructions with sequencing and transcriptome analysis to gain further detail of immune profiles.

### End of study definition

The study will end after the last participant completes visit 6 and all stored samples have been analysed.

### Data management

We will follow the NHS Code of Confidentiality. Data will be handled in accordance with GDPR and DPA 2018. Pseudoanonymisation of personal information will be used for all data entry, research output and electronic sharing of information between the research team. The pseudoanonymisation code will be kept in the site file.

Personal information will only be used on the consent form and the pseudoanonymisation code. Personal information will be used to contact participants if needed and to remind participants of study dates. Personal study data (paper records) will be stored/accessed for 12 months – 3 years after the study has ended. These will be kept within a locked cupboard within a locked office or clinical room.

Fully anonymised data will be shared with fellow researchers via conference presentation and via publication of the results in scientific journals. Data will be shared between the research team named in the application.

All digital storage devices are password locked. Information sent electronically will be encrypted. Anonymisation codes and case-report files will be kept in separate locked cupboards at Barts and the London Faculty of Medicine and Dentistry.

Routine biochemistry will be measured in Barts Health NHS Trust clinical laboratory by the lab biochemists.

### Samples size determination

Our previous evidence indicated that at 72h following cantharidin application 64% (9/14) of blisters had resolved in women whilst 0% (0/13) had resolved in age-matched men (17). If we assume for this study a similar difference for contingency analysis using Fisher’s Exact test with α=0.05 12 volunteers in each group are required to give 90% power to detect this difference between the sexes at this timepoint. If we take a conservative estimate of only 40% resolved in women at the 48h timepoint then with 15 volunteers in each group, we achieve a power of 80%. To account for dropout or accidental blister bursting an additional 2 volunteers in each group will be recruited. Thus, to ensure sufficient power to detect differences at both timepoints where resolution may occur 17 volunteers in each group will be recruited with a total of 34 volunteers. No resolution is expected at 24h.

### Statistical analysis

The number of participants with blister resolution at each timepoint (recorded as the absence of the blister for that relevant timepoint) will be described as a number and percentage for each sex. The relevant effect size is the difference in proportion between sexes. Fisher’s exact tests will compare blister resolution between sexes at each timepoint.

Blister volume for all blisters will be calculated with resolved blisters given a volume of 0mL. The average blister volume at each timepoint will be described as median (interquartile range) for each sex. A Two-way ANOVA will compare blister volume between sexes across each timepoint. Post-hoc testing will focus on comparing the groups at each timepoint.

The total number of inflammatory cells in the blister fluid will be calculated with resolved blisters given a value of 0 cells. The average number of inflammatory cells at each timepoint will be described as median (interquartile range) for each sex. A Two-way ANOVA will compare the number of inflammatory cells between sexes across each timepoint. Post-hoc testing will focus on comparing the groups at each timepoint. Blister volume and total number of inflammatory cells will correlate with each other due to the nature of blister size. To account for this an additional normalised interpretation of number of inflammatory cells will also be analysed and this is the number of inflammatory cells per ml of blister fluid.

Two-way ANOVA will also be used to compare secondary outcomes between the groups including leukocyte subsets, concentration of lactate and LDH, numbers of apoptotic and dead cells and markers of blister efferocytosis. Post-hoc testing will focus on comparing the groups at each timepoint.

### Ethical considerations

The study protocol, participant facing documents, recruitment material and amendments were submitted by the chief investigator to the NHS Health Research Authority REC (Yorkshire & The Humber - Bradford Leeds Research Ethics Committee, reference 22/YH/0244). Prior to the commencement of recruitment, written approval from the REC and sponsor (Queen Mary University of London) was obtained.

### Safety considerations

The risks of this study are negligible. Venepuncture may cause mild bruising/pain and will only be carried out by experienced individuals. Cantharidin may cause some mild discomfort whilst the blister is forming, all care will be taken to avoid removing the flaccid, skin cap as healing is aided by its presence. In previous studies using this technique, there has been no permanent sequelae, such as scars and any pigmentation has usually disappeared by 26 weeks. However, our recent experience in a pilot study suggests that the recovery time in non-Caucasians can be prolonged (up to 4 months) and thus we will ensure all volunteers understand this issue prior to giving consent. Our previous evidence indicates that the recovery time from burst blisters is substantially longer. Once the cantharidin is applied the area will be dressed, the blisters protected and volunteers will be provided with guidance on how to protect the blisters from bursting.

### Safety reporting

Serious Adverse Events (SAEs) that are considered to be ‘unexpected’ and ‘related’ will be reported to the sponsor within 24 hours of learning of the event and to the REC within 15 days in line with the required timeframe. The treatment code for the participant will be broken when reporting an ‘unexpected and related’ SAE. The unblinding of individual participants by the PI / CI in the course of a clinical study will only be performed if necessary for the safety of the study participant.

### Patient and public involvement

The study protocol underwent independent peer review via the William Harvey Research Peer Review Committee and by the Queen Mary University of London and Barts NHS Trust Joint Research Management Office. Study documents were also reviewed by the NHS Health Research Authority REC which includes lay members.

### Dissemination

All relevant data from this study will be submitted to peer review journals for publication following the termination of the study in line with sponsor trust publication policy. Study participants will be asked if they wish to be emailed a short summary of the study findings when available.

## Discussion

In this comparative study we aim to understand the molecular pathways involved in inflammatory resolution in females, compared to males, using a cantharidin blister model of acute inflammation. This study will offer novel insights into sex differences in inflammatory resolution pathways and will form the basis for a larger study in patients with CAD aimed at understanding whether this process is impaired in females with CAD and whether deficient resolution may underlie their susceptibility to worse outcomes following ACS. Ultimately, we hope that this will allow us to identify potential novel therapeutic targets implicated in inflammatory resolution in CAD, as well as leading to a greater understanding of sex differences in health and disease.

### Trial Status

The current protocol version is 1.7 and has been subject to several minor amendments. The original approved protocol 1.2 was subject to a minor amendment on 14/12/2023 to allow photographs to be taken of the blisters. The protocol was subject to an additional minor amendment on 3/2/2023 to provide more detail of sample analysis and add current smoking as a formal exclusion criterion. The protocol was further amended on 29/11/2023 to recruit a further 3 healthy volunteers to account for data loss. Finally, the protocol was amended on 17/6/2024 to provide the correct study duration. All amendments were submitted to and reviewed by the sponsor, when implemented the local research team was informed of changes. The first participant was recruited and included in December 2022 and recruitment was completed in January 2024. Sample analysis and corresponding data collection is currently ongoing.

## Supporting information

Spirit checklist

WHO trial registration data set

## Data Availability

As this is a protocol paper no data was generated or analysed as part of this manuscript.

## Supplementary information

Resolve-Sex Statistical Analysis Plan v1.1

Resolve-Sex Participant Information Sheet v1.5

Resolve-Sex Informed Consent Form v1.1

Resolve-Sex Spirit Checklist

Resolve-Sex WHO trial registration data set

## Administrative information

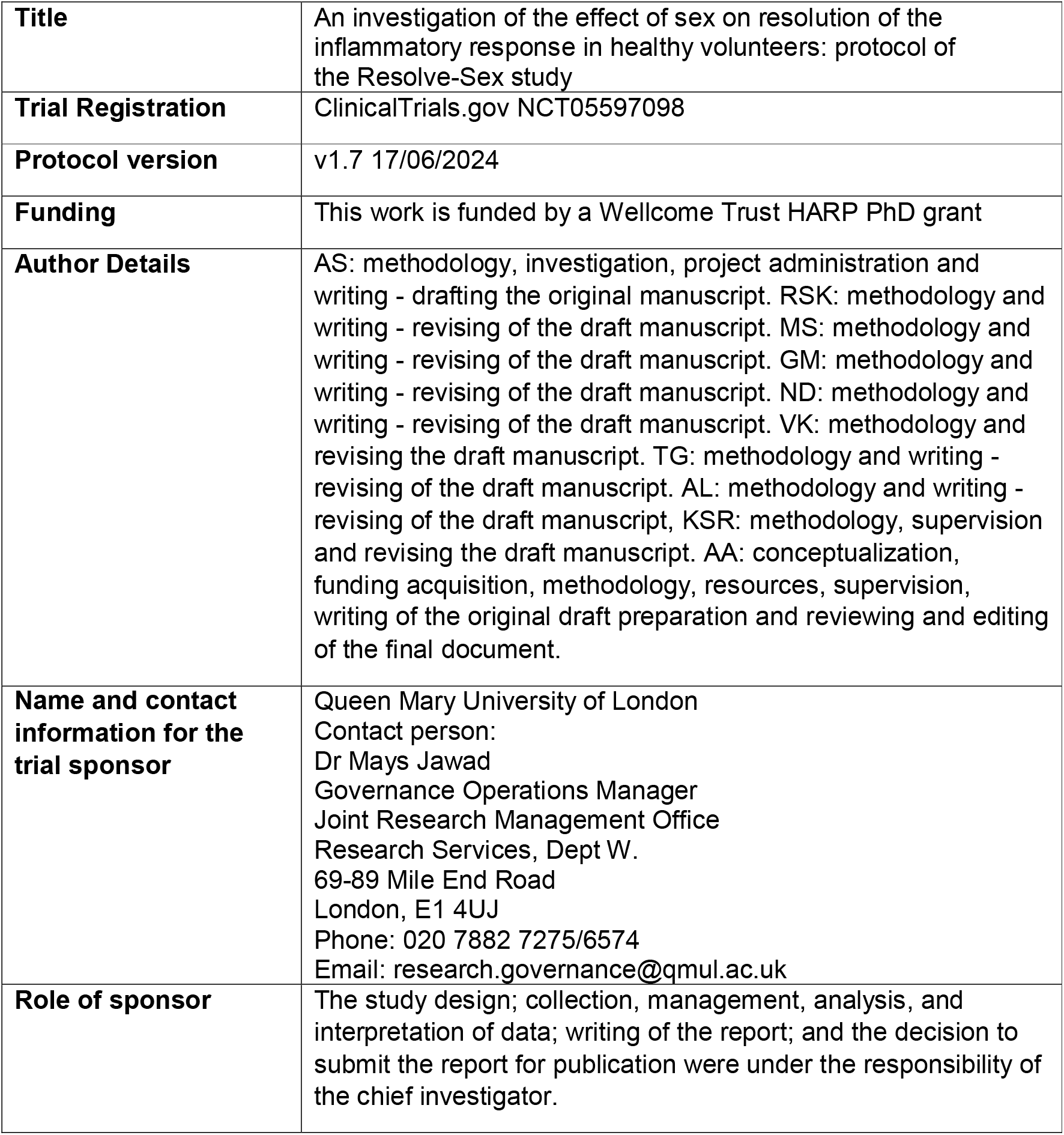

## Acknowledgements

All authors listed above fulfil all three International Committee of Medical Journal Editors (ICMJE) guidelines for authorship, which are 1) substantial contributions to conception and design, acquisition of data or analysis and interpretation of data; 2) drafting the article or revising it critically for important intellectual content; and 3) final approval of the version to be published. AS: methodology, investigation, project administration and writing - drafting the original manuscript. RSK: methodology and writing - revising of the draft manuscript. MS: methodology and writing – revising of the draft manuscript. GM: methodology and writing— revising of the draft manuscript. ND: methodology and writing - revising of the draft manuscript. VK: methodology and revising the draft manuscript. TG: methodology and writing - revising of the draft manuscript. AL: methodology and writing - revising of the draft manuscript, KSR: methodology, supervision and revising the draft manuscript. AA: conceptualization, funding acquisition, methodology, resources, supervision, writing of the original draft preparation and reviewing and editing of the final document. All authors approved the final version of this paper for submission. This study is supported by the CVCTU, a branch of the Barts CTU UKCRC reg no. 4.

## Declarations

### Ethics approval and consent to participate

The Queen Mary and Barts Trust Joint Research Management Office (JRMO) reviewed this protocol and any amendments prior to submission to the REC. The study protocol and any subsequent amendments, along with materials provided to participants and advertising material, were submitted to Yorkshire & The Humber - Bradford Leeds Research Ethics Committee. The study is performed in agreement with the Declaration of Helsinki and is approved by the REC (Yorkshire & The Humber - Bradford Leeds REC, reference 22/YH/0244). Written, informed consent to participate has been and will continue to be obtained from all participants by appropriately trained study team members according to the delegation log. Consent will be acquired for the collection of biological samples to perform exploratory analyses. Biological samples will be stored in − 80° freezers, in secured units.

### Consent for publication

Not applicable

