## Supplementary material for "An investigation of the effect of sex on resolution of the inflammatory response in healthy volunteers: protocol of the Resolve-Sex study": Spirit checklist

SPIRIT 2013 Checklist: Recommended items to address in a clinical trial protocol and related documents*

| **Section/item** | **Item No** | **Description** | **Addressed on page number** |
| --- | --- | --- | --- |
| **Administrative information** | | |  |
| Title | 1 | Descriptive title identifying the study design, population, interventions, and, if applicable, trial acronym | Page 1 – comparative single centre study |
| Trial registration | 2a | Trial identifier and registry name. If not yet registered, name of intended registry | Page 2 - ClinicalTrials.gov NCT05597098 |
|  | 2b | All items from the World Health Organization Trial Registration Data Set | All items are provided in the supplementary material. |
| Protocol version | 3 | Date and version identifier | Page 11 – see trial status section |
| Funding | 4 | Sources and types of financial, material, and other support | Page 12 – see funding section |
| Roles and responsibilities | 5a | Names, affiliations, and roles of protocol contributors | Page 1 |
|  | 5b | Name and contact information for the trial sponsor | Page 3 |
|  | 5c | Role of study sponsor and funders, if any, in study design; collection, management, analysis, and interpretation of data; writing of the report; and the decision to submit the report for publication, including whether they will have ultimate authority over any of these activities | Page 12 – the study sponsor and funders were not involved in study design, collection, management, analysis, interpretation of data, writing of report and decision to submit. |
|  | 5d | Composition, roles, and responsibilities of the coordinating centre, steering committee, endpoint adjudication committee, data management team, and other individuals or groups overseeing the trial, if applicable (see Item 21a for data monitoring committee) | N/A - As this was a healthy volunteer non-CTIMP study with a relatively small sample size no TSC or DMC was required. Trial management was performed by the research team with the PI having main responsibility. General oversight was provided by the study sponsor. |
| **Introduction** |  |  |  |
| Background and rationale | 6a | Description of research question and justification for undertaking the trial, including summary of relevant studies (published and unpublished) examining benefits and harms for each intervention | Page 4 – outlines the research question and justification along with current literature |
|  | 6b | Explanation for choice of comparators | Page 4 – outlines background literature behind choice of comparators and sex differences |
| Objectives | 7 | Specific objectives or hypotheses | Page 5 – see trial objective |
| Trial design | 8 | Description of trial design including type of trial (eg, parallel group, crossover, factorial, single group), allocation ratio, and framework (eg, superiority, equivalence, noninferiority, exploratory) | Page 6 – see study design |
| **Methods: Participants, interventions, and outcomes** | | |  |
| Study setting | 9 | Description of study settings (eg, community clinic, academic hospital) and list of countries where data will be collected. Reference to where list of study sites can be obtained | Page 5 - see participant selection |
| Eligibility criteria | 10 | Inclusion and exclusion criteria for participants. If applicable, eligibility criteria for study centres and individuals who will perform the interventions (eg, surgeons, psychotherapists) | Page 5 |
| Interventions | 11a | Interventions for each group with sufficient detail to allow replication, including how and when they will be administered | Page 6 – study design |
|  | 11b | Criteria for discontinuing or modifying allocated interventions for a given trial participant (eg, drug dose change in response to harms, participant request, or improving/worsening disease) | Page 9 – see safety consideration. No specific criteria for discontinuing or modifying allocated interventions were required based on previous experience of cantharidin use. |
|  | 11c | Strategies to improve adherence to intervention protocols, and any procedures for monitoring adherence (eg, drug tablet return, laboratory tests) | N/A – application of cantharidin was performed by the research team therefore no adherence monitoring was required. |
|  | 11d | Relevant concomitant care and interventions that are permitted or prohibited during the trial | Page 6 – “Following randomisation, the participant is still eligible to undergo usual care should they require it during the study including concomitant planned and unplanned medical care (unrelated to the study itself) and medication for the time period of physical visits and biological sampling; such an event would exclude them from the study.” |
| Outcomes | 12 | Primary, secondary, and other outcomes, including the specific measurement variable (eg, systolic blood pressure), analysis metric (eg, change from baseline, final value, time to event), method of aggregation (eg, median, proportion), and time point for each outcome. Explanation of the clinical relevance of chosen efficacy and harm outcomes is strongly recommended | Page 5 |
| Participant timeline | 13 | Time schedule of enrolment, interventions (including any run-ins and washouts), assessments, and visits for participants. A schematic diagram is highly recommended (see Figure) | Page 6 – see study design |
| Sample size | 14 | Estimated number of participants needed to achieve study objectives and how it was determined, including clinical and statistical assumptions supporting any sample size calculations | Page 8 – see sample size determination |
| Recruitment | 15 | Strategies for achieving adequate participant enrolment to reach target sample size | Page 5 – see participant selection |
| **Methods: Assignment of interventions (for controlled trials)** | | |  |
| Allocation: |  |  |  |
| Sequence generation | 16a | Method of generating the allocation sequence (eg, computer-generated random numbers), and list of any factors for stratification. To reduce predictability of a random sequence, details of any planned restriction (eg, blocking) should be provided in a separate document that is unavailable to those who enrol participants or assign interventions | Page 6 – see randomisation and blinding process |
| Allocation concealment mechanism | 16b | Mechanism of implementing the allocation sequence (eg, central telephone; sequentially numbered, opaque, sealed envelopes), describing any steps to conceal the sequence until interventions are assigned | Page 6 – see randomisation an blinding process |
| Implementation | 16c | Who will generate the allocation sequence, who will enrol participants, and who will assign participants to interventions | Page 6 – see randomisation and blinding process |
| Blinding (masking) | 17a | Who will be blinded after assignment to interventions (eg, trial participants, care providers, outcome assessors, data analysts), and how | Page 6 – see randomisation and blinding process |
|  | 17b | If blinded, circumstances under which unblinding is permissible, and procedure for revealing a participant’s allocated intervention during the trial | N/A – the study cannot be blind since group allocation is pre-determined based on participant sex. |
| **Methods: Data collection, management, and analysis** | | |  |
| Data collection methods | 18a | Plans for assessment and collection of outcome, baseline, and other trial data, including any related processes to promote data quality (eg, duplicate measurements, training of assessors) and a description of study instruments (eg, questionnaires, laboratory tests) along with their reliability and validity, if known. Reference to where data collection forms can be found, if not in the protocol | Page 7-8 – see methods to be used |
|  | 18b | Plans to promote participant retention and complete follow-up, including list of any outcome data to be collected for participants who discontinue or deviate from intervention protocols | Page 9 – “To account for dropout or accidental blister bursting an additional 2 volunteers in each group will be recruited. Thus, to ensure sufficient power to detect differences at both timepoints where resolution may occur 17 volunteers in each group will be recruited with a total of 34 volunteers.” |
| Data management | 19 | Plans for data entry, coding, security, and storage, including any related processes to promote data quality (eg, double data entry; range checks for data values). Reference to where details of data management procedures can be found, if not in the protocol | Page 8 – see data management |
| Statistical methods | 20a | Statistical methods for analysing primary and secondary outcomes. Reference to where other details of the statistical analysis plan can be found, if not in the protocol | Page 9 – see statistical analysis |
|  | 20b | Methods for any additional analyses (eg, subgroup and adjusted analyses) | Page 9 – see statistical analysis |
|  | 20c | Definition of analysis population relating to protocol non-adherence (eg, as randomised analysis), and any statistical methods to handle missing data (eg, multiple imputation) | N/A – as the intervention is applied by research team there is no specific non-adherence in this study. Missing data will be left as missing as imputation is not feasible. Resolved blisters will be given a value of 0 cells (page 9). |
| **Methods: Monitoring** | | |  |
| Data monitoring | 21a | Composition of data monitoring committee (DMC); summary of its role and reporting structure; statement of whether it is independent from the sponsor and competing interests; and reference to where further details about its charter can be found, if not in the protocol. Alternatively, an explanation of why a DMC is not needed | As this was a unblinded healthy volunteer non-CTIMP study with a relatively small sample size no TSC or DMC was required. Data management was undertaken by the research team with the PI having main responsibility. General oversight was provided by the study sponsor. |
|  | 21b | Description of any interim analyses and stopping guidelines, including who will have access to these interim results and make the final decision to terminate the trial | N/A – an interim analysis will not take place as this is a small healthy volunteer mechanistic study. |
| Harms | 22 | Plans for collecting, assessing, reporting, and managing solicited and spontaneously reported adverse events and other unintended effects of trial interventions or trial conduct | Page 10 – see safety reporting |
| Auditing | 23 | Frequency and procedures for auditing trial conduct, if any, and whether the process will be independent from investigators and the sponsor | N/A no specific audits of trial conduct are planned. The sponsor retains the right to conduct an audit at any time. |
| **Ethics and dissemination** | | |  |
| Research ethics approval | 24 | Plans for seeking research ethics committee/institutional review board (REC/IRB) approval | Page 9 – see ethical considerations |
| Protocol amendments | 25 | Plans for communicating important protocol modifications (eg, changes to eligibility criteria, outcomes, analyses) to relevant parties (eg, investigators, REC/IRBs, trial participants, trial registries, journals, regulators) | Page 11- see trial status |
| Consent or assent | 26a | Who will obtain informed consent or assent from potential trial participants or authorised surrogates, and how (see Item 32) | Page 6 – see study design |
|  | 26b | Additional consent provisions for collection and use of participant data and biological specimens in ancillary studies, if applicable | Page 12 – see ethical approval and consent to participate |
| Confidentiality | 27 | How personal information about potential and enrolled participants will be collected, shared, and maintained in order to protect confidentiality before, during, and after the trial | Page 8 – see data management |
| Declaration of interests | 28 | Financial and other competing interests for principal investigators for the overall trial and each study site | Page 12 – see competing interests |
| Access to data | 29 | Statement of who will have access to the final trial dataset, and disclosure of contractual agreements that limit such access for investigators | Page 8 – see data management |
| Ancillary and post-trial care | 30 | Provisions, if any, for ancillary and post-trial care, and for compensation to those who suffer harm from trial participation | Page 6 – see study design |
| Dissemination policy | 31a | Plans for investigators and sponsor to communicate trial results to participants, healthcare professionals, the public, and other relevant groups (eg, via publication, reporting in results databases, or other data sharing arrangements), including any publication restrictions | Page 10 – see dissemination |
|  | 31b | Authorship eligibility guidelines and any intended use of professional writers | Page 12 – see acknowledgements |
|  | 31c | Plans, if any, for granting public access to the full protocol, participant-level dataset, and statistical code | N/A – we aim to publish the study protocol |
| **Appendices** |  |  |  |
| Informed consent materials | 32 | Model consent form and other related documentation given to participants and authorised surrogates | Supplementary material |
| Biological specimens | 33 | Plans for collection, laboratory evaluation, and storage of biological specimens for genetic or molecular analysis in the current trial and for future use in ancillary studies, if applicable | Page 7 & 12 – methods outlined for analysis |

*It is strongly recommended that this checklist be read in conjunction with the SPIRIT 2013 Explanation & Elaboration for important clarification on the items. Amendments to the protocol should be tracked and dated. The SPIRIT checklist is copyrighted by the SPIRIT Group under the Creative Commons “[Attribution-NonCommercial-NoDerivs 3.0 Unported](http://www.creativecommons.org/licenses/by-nc-nd/3.0/)” license.
