## Supplementary material for "An investigation of the effect of sex on resolution of the inflammatory response in healthy volunteers: protocol of the Resolve-Sex study": WHO trial registration data set

| Primary Registry and Trial Identifying Number | NCT05597098 |
| --- | --- |
| Date of Registration in Primary Registry | 24/10/2022 |
| Secondary Identifying Numbers | IRAS Number: 318684  EDGE number: 153010  REC reference: 22/YH/0244 |
| Source(s) of Monetary or Material Support | This work and AS are funded by a Wellcome Trust HARP PhD grant. KSR was previously funded by a NIHR Clinical Academic Lectureship. |
| Primary Sponsor | Queen Mary University of London |
| Secondary Sponsor(s) | N/A – no secondary sponsor |
| Contact for Public Queries | Amrita Ahluwalia  0207 882 8377  |
| Contact for Scientific Queries | Amrita Ahluwalia  0207 882 8377  |
| Public Title | Resolve-Sex: Investigation of the Distinct Mechanisms Involved in Inflammatory Resolution Between Healthy Men and Women |
| Scientific Title | Resolve-Sex: An investigation of the effect of sex on resolution of the inflammatory response in healthy volunteers |
| Countries of Recruitment | United Kingdom |
| Health Condition(s) or Problem(s) Studied | Inflammation resolution |
| Intervention(s) | Cantharidin  0.1% cantharidin solution in acetone from 0.7% stock solution of cantharone is prepared and applied to a disc of filter paper on the forewarm immediately. 10 μl of cantharidin per disc.  Other Names:  Cantharone 0.1%  The participant will initially attend either in person or virtually for a screening visit for eligibility. Cantharidin will be applied on the second visit to the forearm, back or abdomen (depending on patient preference) via 1cm2 cantharidin soaked filter paper. The participant will then attend for two further cantharidin applications (24 hr and 48 hr after the first application). 72 hours after initial cantharidin application the blister fluid will be collected. Male and female participants will be recruited. |
| Key Inclusion and Exclusion Criteria | Inclusion Criteria  Healthy male and female volunteers.  Aged 18-45.  Volunteers who are willing to sign the consent form.  Exclusion Criteria  Healthy subjects unwilling to consent.  Pregnant, or any possibility that a subject may be pregnant unless in the latter case a pregnancy test is performed with a negative result.  Current breast feeding.  History of any serious illnesses, including recent infections or trauma.  Subjects taking systemic medication (other than contraceptives).  Subjects with recent (2 weeks) or current antibiotic use.  Current smoking. |
| Study Type | Comparative mechanistic single centre study |
| Date of First Enrollment | 12/12/2022 |
| Sample Size | 34 participants |
| Recruitment Status | Complete |
| Primary Outcome(s) | Primary Endpoints  1. Comparison of the numbers of volunteers with a blister present at each timepoint over 24-72h between the sexes.  2. Comparison of blister volume at each timepoint over 24-72h between the sexes.  3. Comparison of blister cell number at each timepoint over 24-72h between the sexes. |
| Key Secondary Outcomes | Secondary endpoints  1. Comparison of blister leucocyte subsets (neutrophil and monocyte) between the sexes at each timepoint.  2. Comparison of blister lactate levels and lactate dehydrogenase (LDH) between the sexes at each timepoint.  3. Comparison of cell death, necrotic and apoptotic cell numbers between the sexes at each timepoint.  4. Comparison of markers of blister efferocytosis between the sexes at each timepoint. |
| Ethics Review | The study was approved by the Yorkshire & The Humber –  Bradford Leeds Research Ethics Committee on 17/10/2022, reference 22/YH/0244. |
| Completion date | N/A - study ongoing |
| Summary Results | N/A – study ongoing |
| IPD sharing statement | No |
